# Association of Opioid Use Disorder With 2016 Presidential Voting Patterns: A Cross-Sectional Study in New York State at Census Tract Level

**DOI:** 10.1101/2020.09.05.20188490

**Authors:** Anthony Xiang, Sina Rashidian, Wei Hou, Richard N Rosenthal, Kayley Abell-Hart, Xia Zhao, Fusheng Wang

**Affiliations:** Department of Applied Mathematics and Statistics, Stony Brook University, Stony Brook, NY; Department of Family, Population and Preventive Medicine, Renaissance School of Medicine, Stony Brook, NY; Department of Computer Science, Stony Brook University, Stony Brook, NY; Department of Biomedical Informatics, Renaissance School of Medicine, Stony Brook, NY; Department of Psychiatry, Renaissance School of Medicine, Stony Brook, NY

**Author notes:** Corresponding Author: Fusheng Wang PhD Department of Biomedical Informatics Department of Computer Science Stony Brook University 2313D Computer Science Stony Brook, NY 11794-8330.

**Keywords:** Opioid Use Disorder, Opioid Poisoning, Racial and Ethnic Disparities, Geographic Variance, Sociodemographic Factors, Presidential Election

## Abstract

**Background:** Opioid overdose related deaths have increased dramatically in recent years. Combating the opioid epidemic requires better understanding of the epidemiology of opioid poisoning (OP) and opioid use disorder (OUD).

**Objective:** We aimed to discover geospatial patterns in problematic opioid use and its correlations with demographic features related to despair and economic hardship, most notably the US presidential voting patterns in 2016 at census tract level in New York State.

**Methods:** This cross-sectional analysis used data from New York Statewide Planning and Research Cooperative System claims data and the presidential voting results of 2016 in New York State from the Harvard Election Data Archive. We included 63,958 patients who had at least one opioid use disorder (OUD) diagnosis between 2010 and 2016, and 36,004 patients with at least one opioid poisoning (OP) diagnosis between 2012 and 2016. A logistic regression model was used to determine the associations between patient level characteristics (sex, age group, race, and payment type) and OUD and OP patient rates at census tract level.

**Results:** Several areas shared similar patterns of OUD rates and Republican vote: census tracts in Western New York, Central New York, and Suffolk County. The Spearman rank correlation between OUD rates and the Republican vote was 0.38 (*P* < 0.0001). A multiple regression model of census tract level demographic and socioeconomic factors explains 29% of the variance in OUD rates, with disability and republican vote the biggest predictors.

**Conclusions:** At the census tract level, opioid use disorder rates were positively correlated with Republican support in the 2016 presidential election, disability, unemployment, and unmarried status. Socioeconomic and demographic features explain a large portion of the association between the Republican vote and opioid use disorder. Together, these findings underscore the importance of socioeconomic interventions in combatting the opioid epidemic.

## INTRODUCTION

The United States is experiencing an epidemic of opioid misuse and abuse, involving both prescribed pain relievers and illegal drugs such as heroin and fentanyl. In 2017, the United States Department of Health and Human Services declared the opioid crisis a public health emergency.^1^ A crucial factor in studying patterns in opioid deaths is geographic variation. Previous studies have shown that certain state or county-level characteristics such as rurality, poverty, educational attainment, health care access, and racial demographics, are associated with higher opioid use.^2–4^

An earlier study observed similarities between geographic variation of opioid use and Republican voters at the county level.^5^ Rather than being directly causal, this association is likely driven by external factors shared by both opioid users and voters for the Republican candidate in the 2016 election. Understanding the nature of this relationship helps to place the opioid epidemic in its larger sociopolitical context, and further illuminates the importance of addressing socioeconomic factors in order to fight the opioid epidemic.

Our aim is to better understand the interconnected relationship between opioid use, Republican voting, and other demographic factors in New York State. Our analysis is at the census tract level, which provides a much higher resolution than previous studies. Census tracts generally contain between 2,500 to 8,000 people,^11^ far fewer than the 100,000-inhabitant average at the county level. This fine-grained analysis makes our spatial correlations much more powerful, better revealing how different factors contribute to opioid use disorder and opioid overdose in communities across New York State.

## METHODS

This study was approved by the Stony Brook University Institutional Review Board and the Office of Quality and Patient Safety, Department of Health of New York State. Informed consent was not needed as the study had no contact with participants, and the data were obtained from a New York State administrative database. The primary research question and analysis plan were not pre-registered on a publicly available platform, and thus the results should be considered exploratory.

### Data collection

The presidential voting results of 2016 were obtained from the Harvard Election Data Archive.^6^ These data provided the number of votes for each candidate at an election precinct level, a geographic region generally smaller than the census tract (CT) level. Several counties (e.g., Wyoming County) had incomplete or incoherent data, so those counties were contacted directly to provide election data. The dataset was joined to a geospatial, precinct level shapefile in ArcGIS Desktop 10.7.1 (Esri, Redlands, CA). The precinct-level voting data was extrapolated to the larger census tract level by area-based estimation (Multimedia Appendix 1). The CT voting counts were a linear combination of the precinct level voting counts and precinct area percentage within that census tract (CT components add up to 1). The number of votes for each candidate was then normalized by US Census population estimates of each census tract.

The demographic data were taken from the American Community Survey (ACS) by the US Census Bureau. Census tract level education, age, marriage, unemployment, income, population, race, gender, disability, and healthcare data (Medicare and Medicaid eligibility) were provided in the 2012 to 2016 ACS 5-year estimates.^7^ These data were mapped to a census tract shapefile. Urban-ness is taken from the 2010 Census Summary File and is calculated as the number of households living in an urban area divided by the total number of households in the CT.^8^

The opioid-related patient information was extracted from the SPARCS database, a central administrative repository for health event claims data for New York State patients.^9^ We extracted patients based on ICD codes (primary and secondary diagnosis codes, ICD-9 from January 1, 2012 to September 30, 2015, and ICD-10 from October 1, 2015 to December 31, 2016). Two cohorts of patients were extracted; first, patients diagnosed with *opioid poisoning* (OP) (Multimedia Appendix 2) between 2012 and 2016, and second, patients diagnosed with *opioid use disorder* (OUD) (Multimedia Appendix 3) between 2010 and 2016. For converting home addresses to geolocations (latitude and longitude), we used EaserGeocoder – an open source geocoding software.^10^ The geocoding process runs in-house, therefore no sharing of patient data is needed. It was not possible to convert all patients’ addresses to geolocations, as some of them were either invalid or PO Box addresses instead of street addresses. These patient geolocations were added to the CT shapefile, then grouped and counted within a census tract. OP and OUD rates per 100,000 persons were calculated for each census tract. The SPARCS data also has patient level demographic and other characteristics, such as gender, age, race, and type of payment.

There are 4919 total CTs in NY State according to ACS 2012–2016 5-year estimates. The 2016 voting data from Harvard Dataverse included data for 4900 (99.6%) CTs. After removing CTs with populations less than 100, 4836 (98.3%) CTs remained. These CTs were then used in the spatial mappings, of which 63 CTs had missing education data, 129 had missing income data, 61 had missing marital/race/gender data, and 69 had missing disability data. Excluding these CTs with missing values left 4777 (97.1%) CTs remaining for CT characteristic analyses.

### Analysis

The analyses were divided into two parts, one at the patient level using the SPARCS dataset and the other at the CT level combining the CT dataset and the SPARCS dataset. First, descriptive statistics of patient level characteristics were calculated for OUD and all other patients in the SPARCS dataset. A logistic regression model was used to determine the associations between patient level characteristics (e.g. sex, age group, race, and payment type) with OUD. Odds ratios and their 95% confidence intervals were estimated based on the logistic regression. Second, maps for crude rates of opioid overdose normalized by population for OP and OUD, and maps for 2016 Republican presidential vote rates were generated for CTs with ArcGIS. The opioid use disorder rates were heavily positively skewed due to the high resolution of the geography level and low counts of opioid use for each CT. Spearman rank order correlations were calculated to evaluate the association between OP/OUD and presidential election voting rates. The averages for census tract level demographics and socioeconomic factors were calculated and compared between the CTs with OUD rates in the lowest (1–25%) quartile, and CTs with OUD rates in the highest (76–100%) quartile using t-tests. To assess the extent to which the Republican presidential vote association with opioid use disorder is explained by CT level characteristics, three regression models were built with the OUD rate as the dependent variable. Model 1 included only the percentage of voting for the Republican presidential candidate. Model 2 adjusted for CT demographic and socioeconomic features, and Model 3 additionally aggregated medical factors and median age. The estimated regression coefficients and partial *R*^2^ were reported. Statistical analyses were performed using R version 3.6.1 (R Foundation for Statistical Computing, Vienna, Austria) and Python 3.7 (Python Software Foundation, Wilmington, DE).

## RESULTS

The patient level characteristics of OUD and all other patients in the SPARCS dataset are shown in Table 1. All patient level characteristics are significantly associated with OUD. Given that a patient has OUD, the patient is 1.735 times more likely to be male than female.

**Table 1:** Characteristics of SPARCS Patients Associated with Opioid Use Disorders in New York State, 2010–2016

| **Characteristic** | **All SPARCS (in and out) patients 2010-16 (N = 210935831)** | **OUD patients 2010-16 (n = 63958)** | **Odds ratio (95% CI)** | **P value** |
| --- | --- | --- | --- | --- |
| **Sex - Female** | 119850432 (56.18%) | 27590 (43.13%) | 1 (Ref) |  |
| Male | 91076257 (43.18%) | 36366 (56.86%) | 1.735 (1.708 1.762) | < .0001 |
| **Age group - <18** | 26893357 (12.75%) | 1713 (2.68%) | 0.117 (0.111, 0.123) | < .0001 |
| 18-24 | 17276865 (8.19%) | 9435 (14.75%) | 1 (Ref) |  |
| 25-39 | 38832961 (18.41%) | 19402 (30.34%) | 0.916 (0.894, 0.939) | < .0001 |
| 40-64 | 74991993 (35.55%) | 24078 (37.65%) | 0.588 (0.575, 0.602) | < .0001 |
| 65+ | 46078300 (21.84%) | 9330 (14.59%) | 0.371 (0.361, 0.382) | < .0001 |
| **Race - White, Non-Hispanic** | 90255132 (42.79%) | 42764 (66.86%) | 1 (Ref) |  |
| Black, Non-Hispanic | 41978134 (19.9%) | 6881 (10.76%) | 0.346 (0.337, 0.355) | < .0001 |
| Hispanic | 50616600 (24.00%) | 9176 (14.35%) | 0.383 (0.374, 0.391) | < .0001 |
| Other | 26489568 (12.56%) | 5097 (7.97%) | 0.406 (0.394, 0.418) | < .0001 |
| **Type of payment - Insurance Company** | 74606242 (35.37%) | 19816 (30.98%) | 1 (Ref) |  |
| Medicare | 43520092 (20.63%) | 14378 (22.48%) | 1.907 (1.867, 1.949) | < .0001 |
| Medicaid | 28390633 (13.46%) | 14564 (22.77%) | 1.260 (1.233, 1.287) | < .0001 |
| Self Pay | 17920248 (8.50%) | 6390 (9.99%) | 1.343 (1.305, 1.381) | < .0001 |
| Other | 28652602 (13.58%) | 8810 (13.77%) | 1.158 (1.129, 1.187) | < .0001 |

Table 2 presents the spearman rank-order coefficients between the Republican presidential vote rate and different opioid use rates. There are positive correlations between the Republican presidential vote and opioid use in all opioid use populations, and the magnitudes of correlations are weak to moderate (correlations ranged from 0.25 to 0.38, all p values < 0.0001). Further, there is a negative correlation between the Democratic presidential vote rate and the OUD diagnosis rate from 2010-2016 (r = −0.35, P < .0001).

**Table 2:**
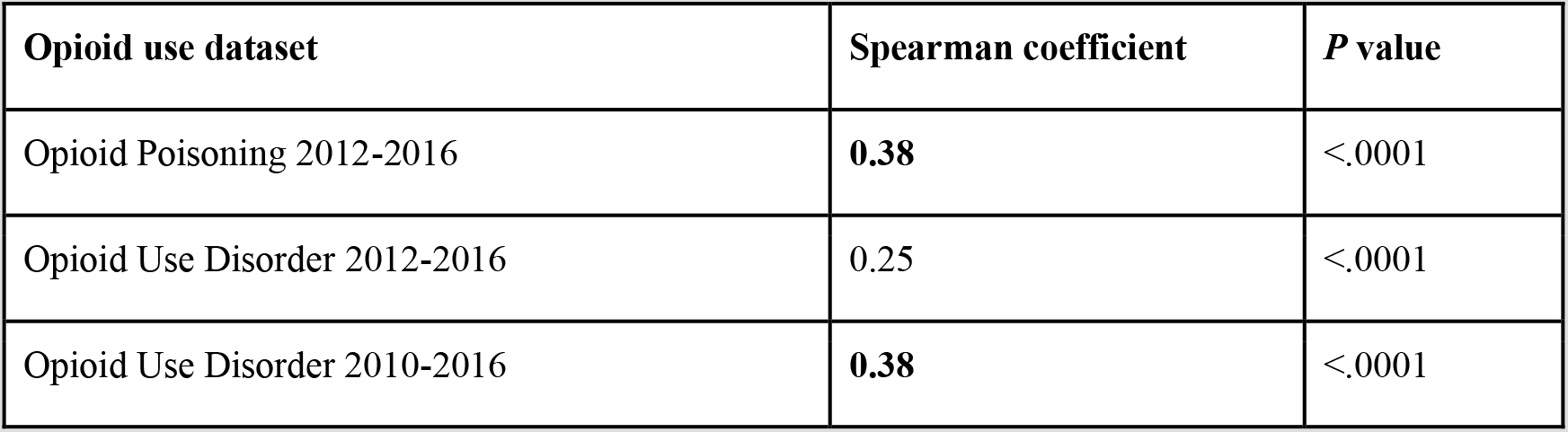
Coefficients of Republican Presidential Vote with Different Opioid Use Patient Populations Opioid use dataset

| Opioid use dataset | Spearman coefficient | $P$ value |
| --- | --- | --- |
| Opioid Poisoning 2012-2016 | **0.38** | $<.0001$ |
| Opioid Use Disorder 2012-2016 | 0.25 | $<.0001$ |
| Opioid Use Disorder 2010-2016 | **0.38** | $<.0001$ |

Figure 1 illustrates opioid use and Republican voting in 4,836 of 4,919 census tracts in New York State. Each map has color ranges ordered by quintiles at the census tract level (exact rates are retracted to comply with CMS cell policy). The first map (A) shows OP rates with a closer look into New York City and Long Island, which have rates among the highest in the state^7^. The areas of higher OP diagnoses were Suffolk County on eastern Long Island, western New York, and Delaware/Broome Counties. Metro areas varied in OP rates. The second map (B) portrays the percentage of the presidential vote for the Republican candidate for each census tract. Note that large urban areas had, for the most part, lower support for the Republican candidate. Several areas shared similar patterns to the OP rates shown in map A: primarily, census tracts in Western New York, Central New York, and Suffolk County. The third map (C) shows OUD rates at the census tract level. The results are similar to map A. The spearman correlation between maps A and B was 0.38 (P < .0001), and the correlation between maps B and C was 0.38 (P < .0001).

**Figure 1.**
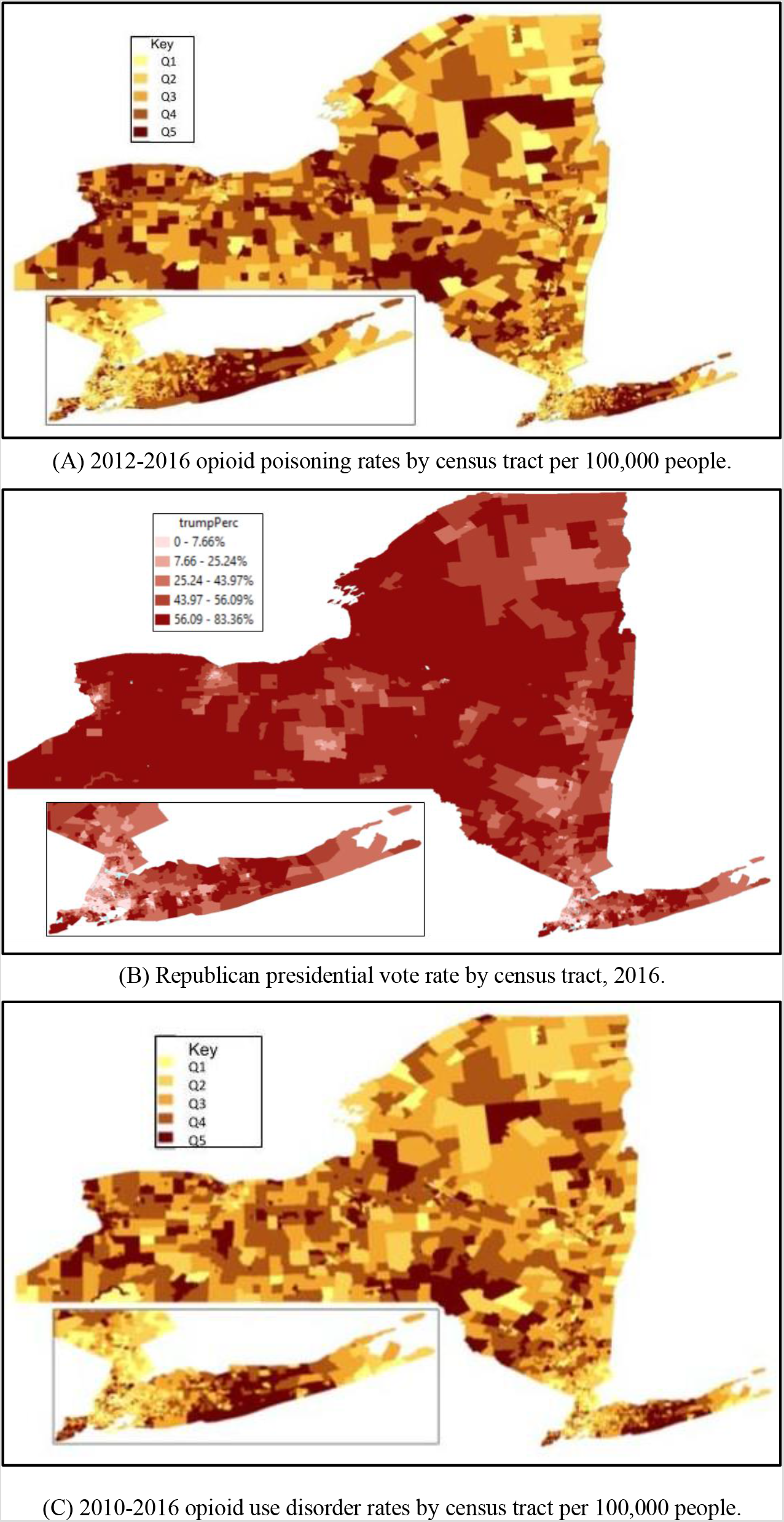
Opioid use and republican presidential vote 2016 in New York State at census tract level.

Next, we examined the CT level characteristics between the Republican presidential vote and opioid use. In Table 3, we tested the differences in the average of various socioeconomic and demographic features at the census tract level between the low OUD and high OUD CTs. CTs were ranked by OUD rates and the lowest and highest quartiles were used for comparison. The Republican presidential vote demonstrated the highest differences between high and low OUD rate census tracts, with the former voting at an average rate of 42.86% (0.56%, P < 0.0001) for the Republican candidate, more than twice the average rate of 20.85%(0.55%) for lower OUD rate census tracts. Other characteristics with relatively large inter-quartile differences include % population with disabilities, % white population, and % of households in urban areas.

**Table 3.**
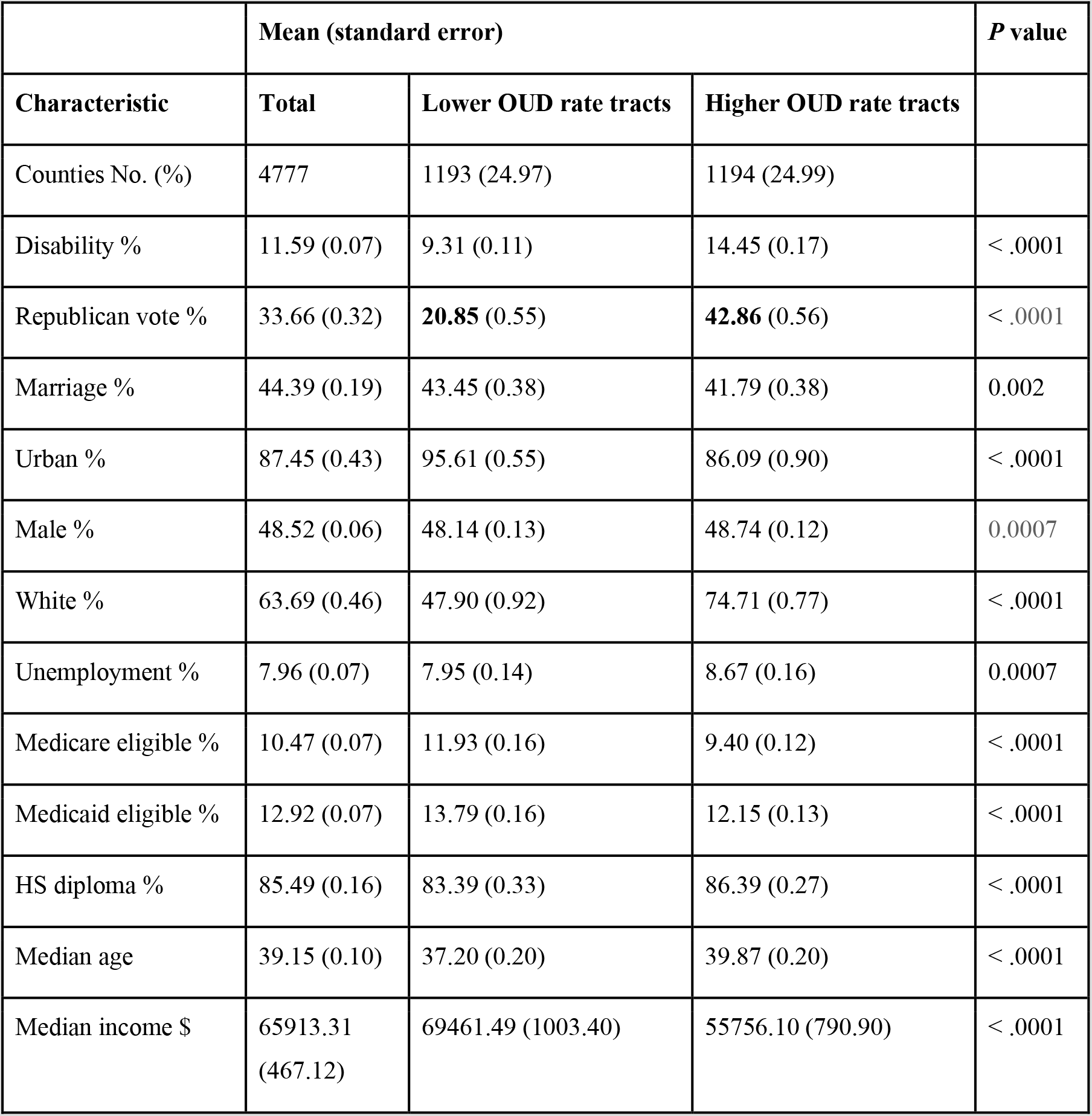
Characteristics Associated with Opioid Use Disorder Rates for Census Tracts with Lower Opioid Use

Finally, we controlled for CT level factors and analyzed the extent to which the Republican presidential vote explains the variation of OUD rates. Table 4 shows 3 regression models, excluding and including the census tract features. Model 1 only contains the Republican vote, which has a positive relationship and explains roughly 5% of the county-level variation in opioid use. Model 2 accounts for several census tract characteristics in addition to the Republican vote, and explains 23% of the variation in OUD rates. The Republican vote still explains 5% of the census tract level variation. Model 3 includes all of the characteristics in Table 3, adding to Model 2 healthcare-related factors (Medicare, Medicaid eligibility, disability) as well as median age. The features explain 29% of the variation in OUD rates and the presidential vote explains 4% of the CT level variation. From models 2 and 3, the most prominent variables that explain the variation in census tract OUD rates are disability rates, percentage of Republican vote, and marriage rates.

**Table 4:** Socioeconomic and Demographic Factors Associating the Republic Vote with Opioid Use Disorder Rates per 100,000 people, 2010–2016

| Characteristic | R<sup>2</sup> | Partial | Regression coefficient | Standard error | P value |
| --- | --- | --- | --- | --- | --- |
| **Model 1** | 0.05 |  |  |  |  |
| Intercept |  |  | 0.137 | 0.005 | < .0001 |
| Republican vote % |  | 0.05 | 0.002 | 0.0001 | < .0001 |
| **Model 2** | 0.23 |  |  |  |  |
| Intercept |  |  | 0.04 | 0.05 | 0.34 |
| Marriage % |  | 0.07 | -0.006 | 0.0003 | < .0001 |
| Republican vote % |  | 0.05 | 0.004 | 0.0002 | < .0001 |
| White % |  | 0.02 | 0.001 | 0.0002 | < .0001 |
| Urban household % |  | 0.01 | 0.0008 | <.0001 | < .0001 |
| Unemployment % |  | 0.01 | 0.005 | 0.0006 | < .0001 |
| Median income, per |  | 0.004 | -0.0005 | 0.0001 | < .0001 |
| \$1000 | | | | | |
| Male % |  | 0.004 | 0.003 | 0.0006 | < .0001 |
| HS diploma % |  | <0.0001 | 0.0002 | 0.0003 | 0.60 |
| **Model 3** | 0.29 |  |  |  |  |
| Intercept |  |  | -0.20 | 0.05 | < .0001 |
| Disability % |  | .05 | 0.01 | 0.0007 | < .0001 |
| Republican vote % |  | .04 | 0.003 | 0.0002 | < .0001 |
| Marriage % |  | .04 | -0.005 | 0.0003 | < .0001 |
| Urban % |  | .02 | 0.001 | <.0001 | < .0001 |
| Male % |  | .01 | 0.004 | 0.0006 | < .0001 |
| White % |  | .008 | 0.0009 | 0.0001 | < .0001 |
| Unemployment % |  | .004 | 0.003 | 0.0006 | < .0001 |
| Medicare eligible |  | .003 | -0.006 | 0.001 | < .0001 |
| Medicaid eligible |  | .002 | 0.004 | 0.001 | 0.006 |
| HS diploma % |  | < .0001 | 0.0002 | 0.0003 | 0.59 |
| Median age |  | < .0001 | 0.002 | 0.0005 | 0.68 |
| Median income \$ | | < .0001 | <.0001 | 0.0001 | 0.96 |

## DISCUSSION

The demographic findings for OUD in NY State were generally consistent with recently published epidemiology of the US opioid epidemic, in that young adult white males are over-represented.^12^

We have explored the specific geographic relationships between opioids, voting patterns, and demographic features like disability and unemployment. Disability may be the easiest factor to explain. In the United States, the largest proportion of years lived with a disability is attributable to chronic non-cancer pain, and globally, musculoskeletal (i.e., back and neck) pain is the third leading cause of disability-adjusted life-years.^13^ As chronic pain is well-described as the most common source of chronic disability in the US, and opioid treatment is also well-described as increasing the odds-ratio for the development of OUD, especially with chronic exposure,^14,15^ it is reasonable to expect that the odds ratio for OUD is increased in patients with chronic disabling conditions.

Next, the small but significant contribution of differences in marriage status is also meaningful in the context of social and economic changes that have paralleled and likely contributed to the arc of the opioid epidemic over the past 25 years.^17^ Monnat and Brown (2017) describe “landscapes of despair”—the small cities and rural areas where over several decades social and family conditions have been deteriorating as economic distress (e.g. job loss due to manufacturing and natural resource industry decline) has been mounting.^18^ They found, consistent with our findings, the highest percentage of 2016 Republican voting over the 2012 baseline in the top quartile of counties with the lowest well-being, which included higher separation/divorce rates as compared to the quartile of counties with the highest well-being.^18^ These locales are also where the 2016 Republican candidate over-performed compared to Republican voting patterns in the 2012 election: counties with the highest rate of deaths of despair, i.e., those with the highest drug, alcohol and suicide mortality rates attributable in large measure to economic distress and a large working class.^19^

Understanding these landscapes of despair is crucial because opioids are an anodyne to both physical and emotional pain. This fact may also help explain the relationship between opioids and voting patterns. A political candidate may appeal to residents of these landscapes by resonating with their emotional needs, and even presenting themself as a kind of anodyne by promising to uplift them both economically and socio-politically.

Although a causal relationship cannot necessarily be inferred, our model clusters increased disability, voting Republican, and lower odds of having a marital partner, with risk for OUD. Our findings highlight the relationship between opioid dependence and factors related to despair, suggesting that socioeconomic growth may be necessary to successfully fight the opioid epidemic, in addition to traditional interventions like improved access to OUD treatment. Disability, unemployment, and non-married status do not have to cause despair, but are likely to do so in communities that lack a safety net, both economically and socially. Understanding and responding to the needs of these “landscapes of despair” may be key to reversing the opioid epidemic, and may also affect the political direction of the United States.

Our study has a few limitations regarding the method and underlying assumptions about the population. It is important to note that the population base containing the sample that voted Republican in 2016 is not the same as the population base data we used to determine opioid use disorder, but rather they were generalized and configured to the census tract level. In addition, for the purposes of constructing our statistical analyses, we assumed in these populations that socioeconomic/demographic factors affect OUD rates. Lastly, in order to converge the datasets appropriately, we assumed that the population in which we drew data to determine OUD was also alive and voting in the 2016 election.

## CONCLUSIONS

The association between the 2016 Republican presidential vote and opioid use disorder highlights the demographic, geographic, and socioeconomic characteristics that underpin both features. Studying opioid use at a finer grain geospatial level provides a unique opportunity for a more precise understanding of the opioid epidemic at large scale.

## Data Availability

There are several data sources used in this study. Patients data is from New York Statewide Planning and Research Cooperative System (SPARCS) which is available for research purposes upon request and approval. The demographic data were taken from the American Community Survey (ACS) by the US Census Bureau. The presidential voting results of 2016 were obtained from the Harvard Election Data Archive. These two datasets are available online.

## ACKNOWLEDGEMENTS

This work was supported in part by National Science Foundation Advanced Cyberinfrastructure no. 1443054 and by National Science Foundation Information and Intelligent Systems 135088. The authors disclose no conflicts of interest.

